# Association of axial length and changes in aqueous depth with refractive outcomes in Chinese primary angle closure glaucoma patients

**DOI:** 10.64898/2026.04.10.26350671

**Authors:** Yaping Yang, Tsz Kin Ng, Junyi Chen, Xinghuai Sun, Li Wang

## Abstract

**Purpose:** To identify the ocular biometric parameters associated with refractive outcomes in Chinese Primary angle closure glaucoma (PACG) patients receiving phacoemulsification and intraocular lens (IOL) implantation (PEI) surgery.

**Methods:** 165 Chinese PACG patients receiving PEI and goniosynechialysis (GSL) and 53 cataract patients as controls only receiving PEI surgery were recruited. The prediction accuracy of IOL power calculation was assessed by the prediction error (PE), mean absolute error (MAE), median absolute error (MedAE), and proportions of eyes with a PE within ± 0.25 diopters (D), ± 0.50 D, ± 0.75 D, and ± 1.00 D. The association of different ocular biometric parameters with the PE of IOL calculation were evaluated.

**Results:** The PACG patients had significantly higher absolute of PE as compared to the control subjects, especially the acute PACG patients. The axial length (AL), changes in aqueous depth pre- and post-surgery (△AD), and the ratio of △AD/AL were significantly associated with the PE in acute PACG patients. The association of △AD with the PE of IOL power calculation was found in PACG patients with AL ≥ 22 mm.

**Conclusions:** This study revealed the association of AL and △AD with the PE of IOL calculation in Chinese PACG patients. Precisely predict the △AD is necessary for acute PACG patients, especially for those with AL ≥ 22 mm, to improve the refractive outcomes.

## Introduction

Glaucoma is a leading cause of irreversible blindness and visual impairment worldwide [1]. Primary angle closure glaucoma (PACG) is a major subtype of primary glaucoma in Asia [2]. Angle closure is characterized by the mechanical contact of the iris onto the trabecular meshwork (TM)-peripheral anterior synechiae (PAS), which in turn leads to the blockage of aqueous outflow and intraocular pressure (IOP) elevation [3]. PACG can be sub-classified into acute PACG (APACG) and chronic PACG (CPACG) [4]. Patients with APACG experience a sudden IOP elevation as the iris rapidly covers the TM, while patients with CPACG experience slow and asymptomatic increase in IOP as the iris gradually covers the TM [5].

Clinical treatments for PACG varies depending on the severity of the angle closure. When the degree of PAS is mild, laser peripheral iridotomy, phacoemulsification alone or in combination with topical IOP-lowering medication can be applied to lower the IOP [6]. When extensive PAS develops, phacoemulsification combined with goniosynechialysis (phaco-GSL) surgery has been reported to be effective for PACG to restore the normal outflow pathway and reduce IOP [7]. However, some studies have reported inferior precision of refractive outcomes in PACG patients after phaco-GSL surgery due to the shallow anterior chamber and lens instability caused by the zonular weakness [8-12]. Although newly developed intraocular lens (IOL) formulas are based on anterior chamber depth (ACD), lens thickness (LT), keratometry (K), and axial length (AL), these variables may be distorted and structurally abnormal in PACG patients. Based on these circumstances, determination of the association of these ocular biometric parameters with refractive outcomes after IOL implantation in PACG patients is warranted. In this study, we evaluated the refractive outcomes in IOL power calculation and analyzed the association of different ocular biometric parameters with the prediction error of refractive outcomes in Chinese PACG patients.

## Methods

### Study subjects

In total, 165 Chinese PACG patients, including 106 APACG (106 eyes) and 59 CPACG patients (59 eyes), and 53 cataract patients as controls (53 eyes) were recruited from the Eye and Ear Nose Throat Hospital, Fudan University, Shanghai, China.

The inclusion criteria for APACG included [13]: (1) a history of at least two typical symptoms (ocular or periocular pain, nausea or vomiting, and/or intermittent blurring of vision with halos); (2) an IOP spike > 30 mmHg; (3) the presence of at least three signs (obvious conjunctival hyperemia, corneal epithelial edema, med-dilated pupil, and/or shallow anterior chamber); (4) Angle closure was confirmed by gonioscopic examination; (5) glaucomatous optic neuropathy or visual field defect. The inclusion criteria for CPACG included: (1) the absence of acute attack symptoms or signs; (2) PAS confirmed by gonioscopy; (3) chronic IOP elevation (> 21 mmHg); and (4) glaucomatous optic neuropathy or visual field defect. The subjects were exclude if they had secondary glaucoma, any other eye disease that could affect visual acuity or the visual field, histories of any type of ocular incisional surgery, ocular trauma or systemic diseases. If both eyes of a patient met the inclusion criteria, only one eye was randomly selected. Both APACG and CPACG patients were initially treated with medications to lower the IOP. All topical ocular medications were continued until the time of the surgery.

All control subjects diagnosed with cataract did not have any other ocular diseases, surgeries and systemic diseases, except mild refractive errors. This study has been approved by the medical ethics committee of the Eye and Ear Nose Throat Hospital, Fudan University, which was in accordance with the tenets of the Declaration of Helsinki. Written informed consents were obtained from all the study subjects after explanation of the nature and possible consequences of the study during the period from January 2023 to March 2025.

### Ophthalmic examinations and intraocular lens power calculation

Comprehensive ophthalmic examinations, including distance-corrected visual acuity (DCVA) in logMAR scale, refractive error, IOP measurement by Goldmann applanation tonometry, slit-lamp examination, fundus examination, and ocular biometric parameter measurement, were performed in all study subjects. Visual field test (OCTOPUS 101 automated perimetry) were performed in all PACG patients. Aqueous depth (AD, measured from corneal endothelium to anterior lens surface), central corneal thickness (CCT) were measured by Lenstar (LS900; Haag-Streit, Köniz, Switzerland). Other ocular biometric parameters, including ACD (measured from corneal epithelium to anterior lens surface), AL, and LT were measured by the IOLMaster700^®^ (Carl Zeiss Meditec AG, Germany) with optimized values. △AD is calculated as the post-surgery ACD subtracting the pre-surgery ACD. ΔAD reflects the real changes of aqueous depth, which is not affected by CCT. The IOL power prediction were calculated by the Barrett Universal II formula. The prediction accuracy of IOL power calculation was assessed by the prediction error (PE), mean absolute error (MAE), median absolute error (MedAE), and proportions of eyes with a PE within ± 0.25 diopters (D), ± 0.50 D, ± 0.75 D, and ± 1.00 D. PE was defined as the spherical equivalent at one month post-surgery subtracting the predicted refraction of the IOL implantion.

### Phacoemulsification surgery

All PACG patients received phacoemulsification, IOL implantation, and goniosynechialysis. Routine PEI was performed after the administration of topical anesthesia, a standard clear-cornea tunneled phacoemulsification procedure, followed by a foldable intraocular lens (including Tecnis PCB00 or ZA9003 IOL, J & J, Santa Ana, CA, USA) was implanted in the capsular bag. Subsequently, GSL was performed by gently pressing the PAS with a chopper against the peripheral edges of the iris until the entire angle was fully open. All control subjects received phacoemulsification and IOL implantation. After surgery, steroids and antibiotics were administered to all patients.

### Statistical analysis

The data were presented as mean ± standard deviation (SD) for continuous variables and as percentages for categorical variables. Continuous variables were compared using one-way analysis of variance with post-hoc Dunn’s correction. Mann-Whitney U test with post-hoc Dunn’s correction were used to compare the differences of absolute error of prediction error between groups. Multivariable linear regression analyses were performed to evaluate the association of different ocular biometric parameters with PE, with adjustment for age and sex. The odds ratios (OR) and β coefficient with 95% confident intervals (C.I.) were estimated. All statistical analyses were conducted by R (version 4.0.1; http://www.rproject.org). *P* < 0.05 was considered as statistically significant.

## Results

### Demographics of the study subjects

In total, 218 study subjects were recruited, including 106 APACG, 59 CPACG, and 53 control subjects (Table 1). The IOP before the surgery of total PACG (22.24 ± 11.81 mmHg, *P* < 0.001), APACG (23.56 ± 13.19 mmHg, *P* < 0.001), and CPACG patients (19.60 ± 7.90 mmHg, *P* = 0.011) were significantly higher than that of the control subjects (14.70 ± 3.25 mmHg). Moreover, the LT of total PACG (5.04 ± 0.34 mm, *P* < 0.001), APACG (5.07 ± 0.33 mm, *P* < 0.001), and CPACG patients (4.98 ± 0.35 mm, *P* < 0.001) were significantly thicker than that of the control subjects (4.58 ± 0.54 mm). The pre-surgery AL, ACD, and AD of total PACG (AL: 22.51 ± 0.83 mm, *P* < 0.001; ACD: 2.17 ± 0.28 mm, *P* < 0.001; AD: 1.67 ± 0.29 mm, *P* < 0.001), APACG (AL: 22.39 ± 0.85 mm, *P* < 0.001; ACD: 2.11 ± 0.29 mm, *P* < 0.001; AD: 1.60 ± 0.29 mm, *P* < 0.001), and CPACG patients (AL: 22.74 ± 0.76 mm, *P* < 0.001; ACD: 2.29 ± 0.22 mm, *P* < 0.001; AD: 1.80 ± 0.25 mm, *P* < 0.001) were significantly shorter than that of the control subjects (AL: 23.45 ± 0.99 mm; ACD: 2.91 ± 0.43, *P* < 0.001; AD: 2.39 ± 0.40 mm). Consistently, the AD/AL ratio of total PACG (0.07 ± 0.01, *P* < 0.001), APACG (0.07 ± 0.01, *P* < 0.001), and CPACG patients (0.08 ± 0.01, *P* < 0.001) were significantly lower than that of the control subjects (0.10 ± 0.02). Notably, significantly more changes in AD after surgery (ΔAD: 1.84 ± 0.52 mm) and the ratio of the changes in AD to AL (ΔAD/AL; 0.08 ± 0.02) were observed in APACG patients as compared to the control (ΔAD: 1.66 ± 0.36 mm, *P* = 0.047; ΔAD/AL: 0.07 ± 0.02, *P =* 0.007), and CPACG groups (ΔAD: 1.63 ± 0.40 mm, *P* = 0.012; ΔAD/AL: 0.07 ± 0.02, *P =* 0.002).

**Table 1.** Ocular biometirc parameters in primary angle-closure glaucoma and control subjects.

|  | **Control**<br>(n=53) | **APACG**<br>(n=106) | **CPACG**<br>(n=59) | **PACG**<br>(n=165) |  |  |  |  |
| --- | --- | --- | --- | --- | --- | --- | --- | --- |
| Parameters | Male (12/22.64%) | Male (19/17.92%) | Male (32/54.24%) | Male (51/30.91%) |  |  |  |  |
|  | Female (41/77.36%) | Female (87/82.08%) | Female (27/45.76%) | Female (114/69.09%) |  |  |  |  |
|  | Mean±SD | Mean±SD | Mean±SD | Mean±SD | **P<sub>1</sub>** | **P<sub>2</sub>** | **P<sub>3</sub>** | **P<sub>4</sub>** |
| Age (years) | 69.9 ± 7.7 | 66.4 ± 8.5 | 64.4 ± 7.9 | 65.8 ± 8.3 | **0.009** | **<0.001** | **0.001** | 0.136 |
| Axial length (AL) | 23.45 ± 0.99 | 22.39 ± 0.85 | 22.74 ± 0.76 | 22.51 ± 0.83 | **<0.0001** | **<0.0001** | **<0.0001** | **0.009** |
| Central Corneal Thickness (CCT) | 540.98 ± 36.32 | 560.81 ± 49.92 | 534.80 ± 25.04 | 552.09 ± 44.85 | **0.004** | 0.425 | 0.091 | **<0.001** |
| Len Thickness (LT) | 4.58 ± 0.54 | 5.07 ± 0.33 | 4.98 ± 0.35 | 5.04 ± 0.34 | **<0.0001** | **<0.0001** | **<0.0001** | 0.093 |
| Intraocular pressure IOP | 14.70 ± 3.25 | 23.56 ± 13.19 | 19.60 ± 7.90 | 22.24 ± 11.81 | **<0.0001** | **0.011** | **<0.0001** | **0.035** |
| Anterior Chamber Depth (ACD) | 2.91 ± 0.43 | 2.11 ± 0.29 | 2.29 ± 0.22 | 2.17 ± 0.28 | **<0.0001** | **<0.0001** | **<0.0001** | **<0.0001** |
| Aqueous depth (AD) | 2.39 ± 0.40 | 1.60 ± 0.29 | 1.80 ± 0.25 | 1.67 ± 0.29 | **<0.0001** | **<0.0001** | **<0.0001** | **<0.001** |
| ΔAD | 1.66 ± 0.36 | 1.84 ± 0.52 | 1.63 ± 0.40 | 1.76 ± 0.49 | **0.047** | 0.709 | 0.267 | **0.012** |
| ΔAD/AL | 0.07 ± 0.02 | 0.08 ± 0.02 | 0.07 ± 0.02 | 0.08 ± 0.02 | **0.007** | 0.828 | 0.061 | **0.002** |
| AD/AL | 0.10 ± 0.02 | 0.07 ± 0.01 | 0.08 ± 0.01 | 0.07 ± 0.01 | **<0.0001** | **<0.0001** | **<0.0001** | **0.002** |

*P*<sub>1</sub>: APACG vs Control; *P*<sub>2</sub>: CPACG vs Control; *P*<sub>3</sub>: PACG vs Control; *P*<sub>4</sub>: APACG vs CPACG; All P values were adjusted for age and sex. P values were in bold face if < 0.05.
APACG: acute primary angle-closure glaucoma; CPACG: chronic primary angle-closure glaucoma; PACG: primary angle closure glaucoma
Anterior Chamber Depth (ACD): Measured from corneal epithelium to anterior lens surface, by IOLMaster700® pre-surgery.
Aqueous depth (AD): Measured from endothelium to anterior lens surface, by Lenstar pre-surgery.
ΔAD: Changes in aqueous depth without CCT, calculated as one month post-surgery ACD subtracting the pre-surgery ACD measured by IOLMaster700®.

### Refractive outcomes in intraocular lens power calculation

For the prediction accuracy of IOL power calculation, the MedAE were larger in total PACG (0.44 D) and APACG groups (0.61 D) as compared to the control (0.29 D) and CPACG groups (0.26 D; Table 2). Consistently, the MAE were also larger in total PACG (0.61 D) and APACG groups (0.71 D) as compared to the control (0.37 D) and CPACG groups (0.43 D). Compared to the control group, the absolute of PE were significantly different in total PACG (*P* = 0.007) and APCAG groups (*P* < 0.001), and significance differences between the APACG and CPACG groups were also observed (*P* = 0.005). Lower proportions of eyes within PE ± 0.50 D were observed in total PACG (55.16%) and APACG groups (46.22%) as compared to the control group (71.69%) and the CPACG group (71.18%). In addition, higher percentage of hyperopic refactive error shift were observed in total PACG, APACG, and CPACG groups as compared to the control group. The propotion of eyes with the absolute of PE > 1.00 D in the APACG group (27.36%) was higher than the control (3.77%) and CPACG groups (11.86%).

**Table 2.**
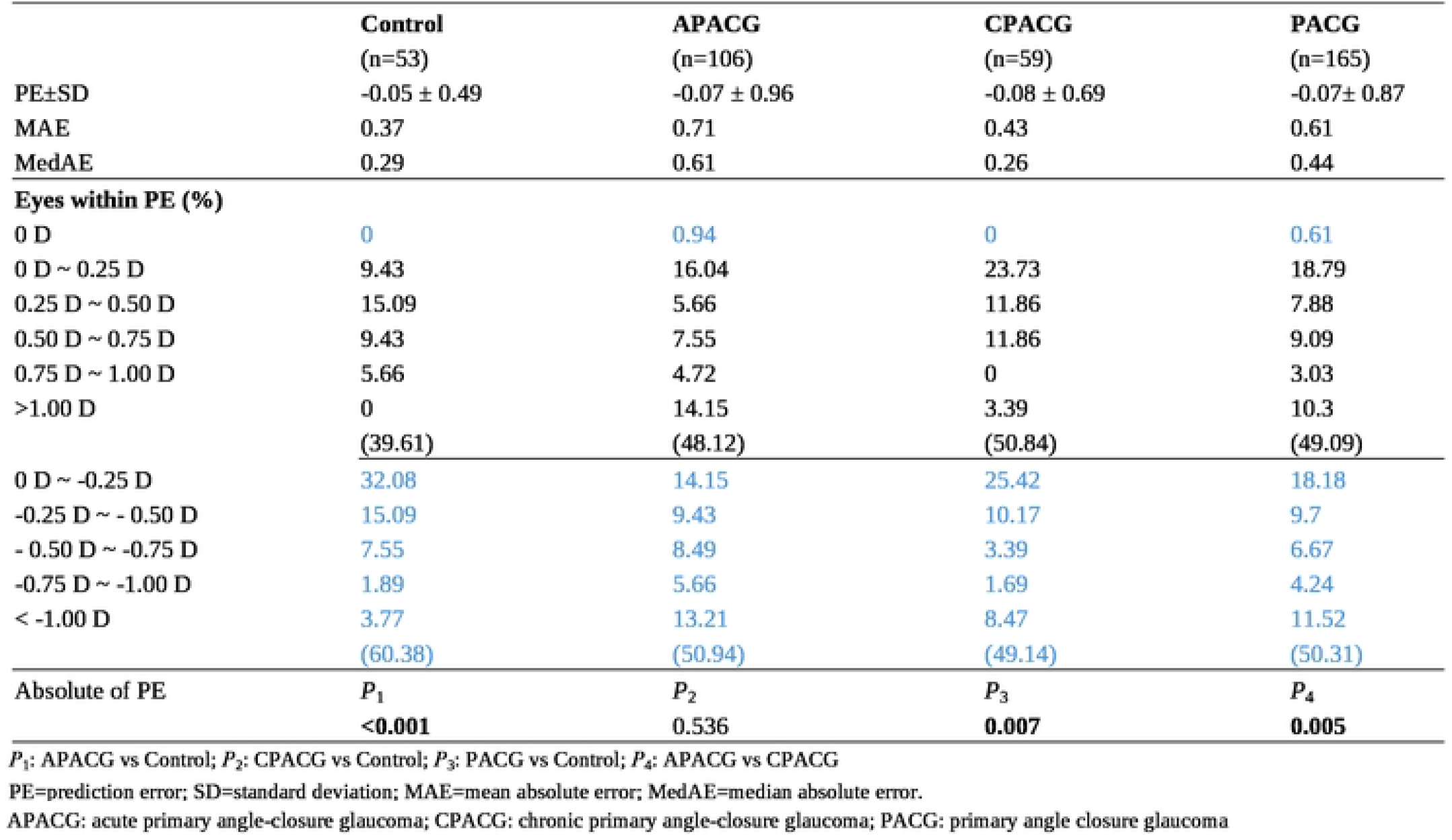
Refractive errors in primary angle-closure glaucoma groups and control subjects.

With the stratification of AL and AD, the PE, MAE, and MedAE were lower in the subjects with AL ≥ 22 mm than those with AL < 22 mm, and significant differences in absolute of PE were found between the two groups (*P* = 0.030; Table 3). The proportions of eyes within PE ± 0.50 D in the subjects with AL ≥ 22 mm (59.2%) were higher than those with AL < 22 mm (44.4%). For the subjects with AL < 22 mm, the proportions of eyes within PE ± 0.50 D in subjects with AD > 2 mm (40.0%) were lower than those with AD ≤ 2 mm (45.0%), but the absolue of PE was not signicant (*P* = 0.617). In contrast, for the subjects with AL ≥ 22 mm, the proportions of eyes within PE ± 0.50 D in the subjects with AD > 2 mm (71.43%) were higher than those with AD ≤ 2 mm (57.55%), and the absolue of PE between the two groups were also not significant (*P* = 0.145).

**Table 3.**
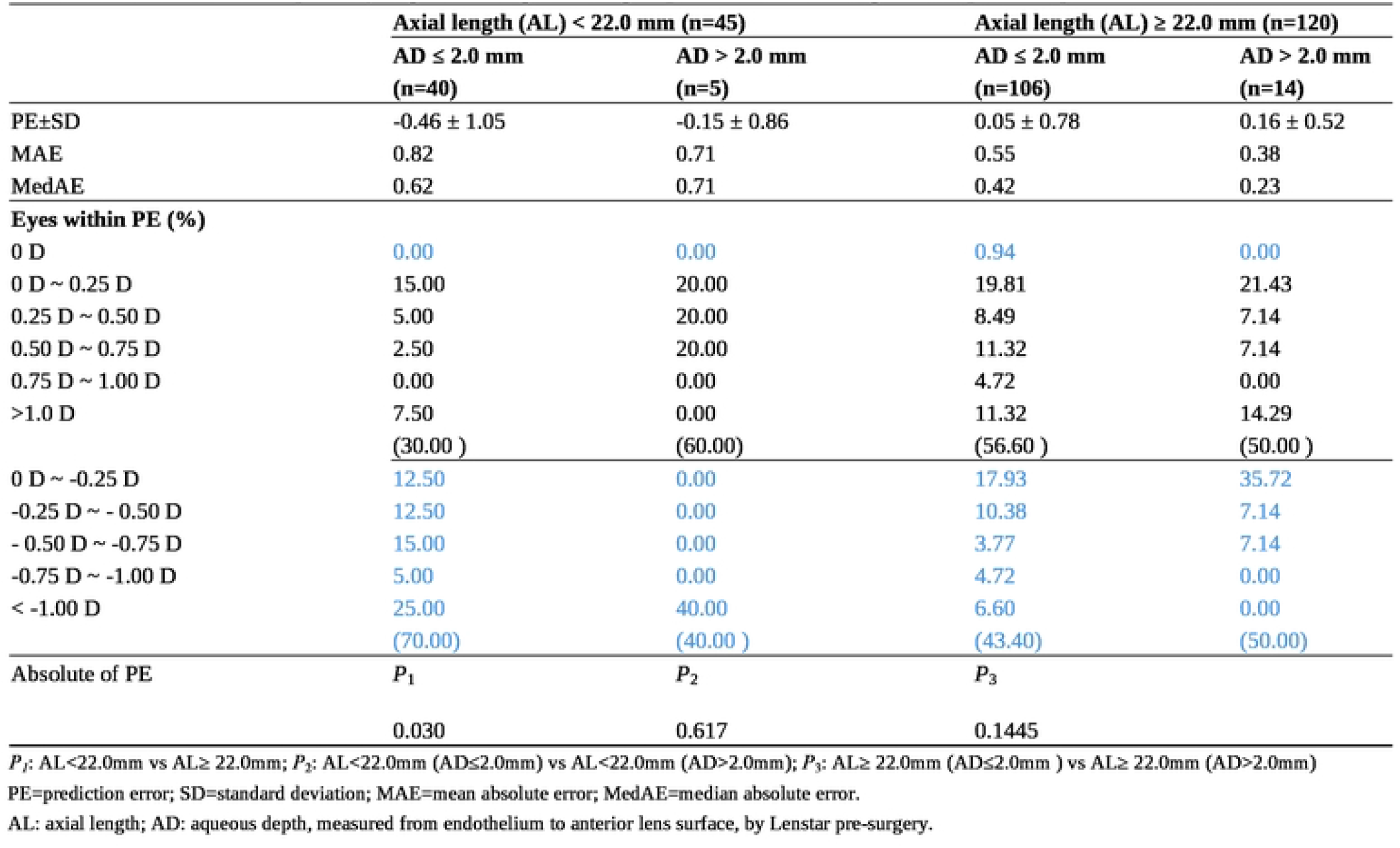
Refractive errors in primary angle-closure glaucoma groups based on axial length and aqueous depth.

|  | Axial length (AL) < 22.0 mm (n=45) |  | Axial length (AL) ≥ 22.0 mm (n=120) |  |
| --- | --- | --- | --- | --- |
|  | AD ≤ 2.0 mm<br>(n=40) | AD > 2.0 mm<br>(n=5) | AD ≤ 2.0 mm<br>(n=106) | AD > 2.0 mm<br>(n=14) |
| PE±SD | -0.46 ± 1.05 | -0.15 ± 0.86 | 0.05 ± 0.78 | 0.16 ± 0.52 |
| MAE | 0.82 | 0.71 | 0.55 | 0.38 |
| MedAE | 0.62 | 0.71 | 0.42 | 0.23 |
| **Eyes within PE (%)** |  |  |  |  |
| 0 D | 0.00 | 0.00 | 0.94 | 0.00 |
| 0 D ~ 0.25 D | 15.00 | 20.00 | 19.81 | 21.43 |
| 0.25 D ~ 0.50 D | 5.00 | 20.00 | 8.49 | 7.14 |
| 0.50 D ~ 0.75 D | 2.50 | 20.00 | 11.32 | 7.14 |
| 0.75 D ~ 1.00 D | 0.00 | 0.00 | 4.72 | 0.00 |
| >1.0 D | 7.50 | 0.00 | 11.32 | 14.29 |
|  | (30.00 ) | (60.00) | (56.60 ) | (50.00 ) |
| 0 D ~ -0.25 D | 12.50 | 0.00 | 17.93 | 35.72 |
| -0.25 D ~ - 0.50 D | 12.50 | 0.00 | 10.38 | 7.14 |
| - 0.50 D ~ -0.75 D | 15.00 | 0.00 | 3.77 | 7.14 |
| -0.75 D ~ -1.00 D | 5.00 | 0.00 | 4.72 | 0.00 |
| < -1.00 D | 25.00 | 40.00 | 6.60 | 0.00 |
|  | (70.00) | (40.00 ) | (43.40) | (50.00) |
| **Absolute of PE** |  |  |  |  |
| | $P_1$ | $P_2$ | $P_3$ | |
|  | 0.030 | 0.617 | 0.1445 |  |

$P_1$ : AL<22.0mm vs AL≥ 22.0mm; $P_2$ : AL<22.0mm (AD≤2.0mm) vs AL<22.0mm (AD>2.0mm); $P_3$ : AL≥ 22.0mm (AD≤2.0mm ) vs AL≥ 22.0mm (AD>2.0mm)
PE=prediction error; SD=standard deviation; MAE=mean absolute error; MedAE=median absolute error.
AL: axial length; AD: aqueous depth, measured from endothelium to anterior lens surface, by Lenstar pre-surgery.

### Association of ocular biometric parameters with prediction error in primary angle closure glaucoma and control subjects

With adjustment for age and sex, multivariable linear regression analysis revealed that preoperative IOP was significantly associated with PE in the PACG patients (β = 0.02, 95% C.I.: 0.00 – 0.03, *P* = 0.018; Table 4). Moreover, AL was significantly associated with PE in total PACG (β = 0.32, 95% C.I.: 0.17 – 0.48, *P* < 0.001) and APACG groups (β = 0.44, 95% C.I.: 0.23 – 0.65, *P* < 0.001). In addition, △AD was significantly associated with PE in total PACG (β = 0.57, 95% C.I.: 0.33 – 0.81, *P* < 0.0001) and APACG groups (β = 0.73, 95% C.I.: 0.40 – 1.05, *P* < 0.0001). Similarly, the △AD/AL ratio was also significantly associated with PE in total PACG (β = 11.80, 95% C.I.: 6.26 – 17.35, *P* < 0.001) and APACG groups (β = 14.87, 95% C.I.: 7.19 – 22.56, *P* < 0.001). However, no significant association was found in LT, AD, and AD/AL with PE in any groups (*P* > 0.05).

**Table 4.**
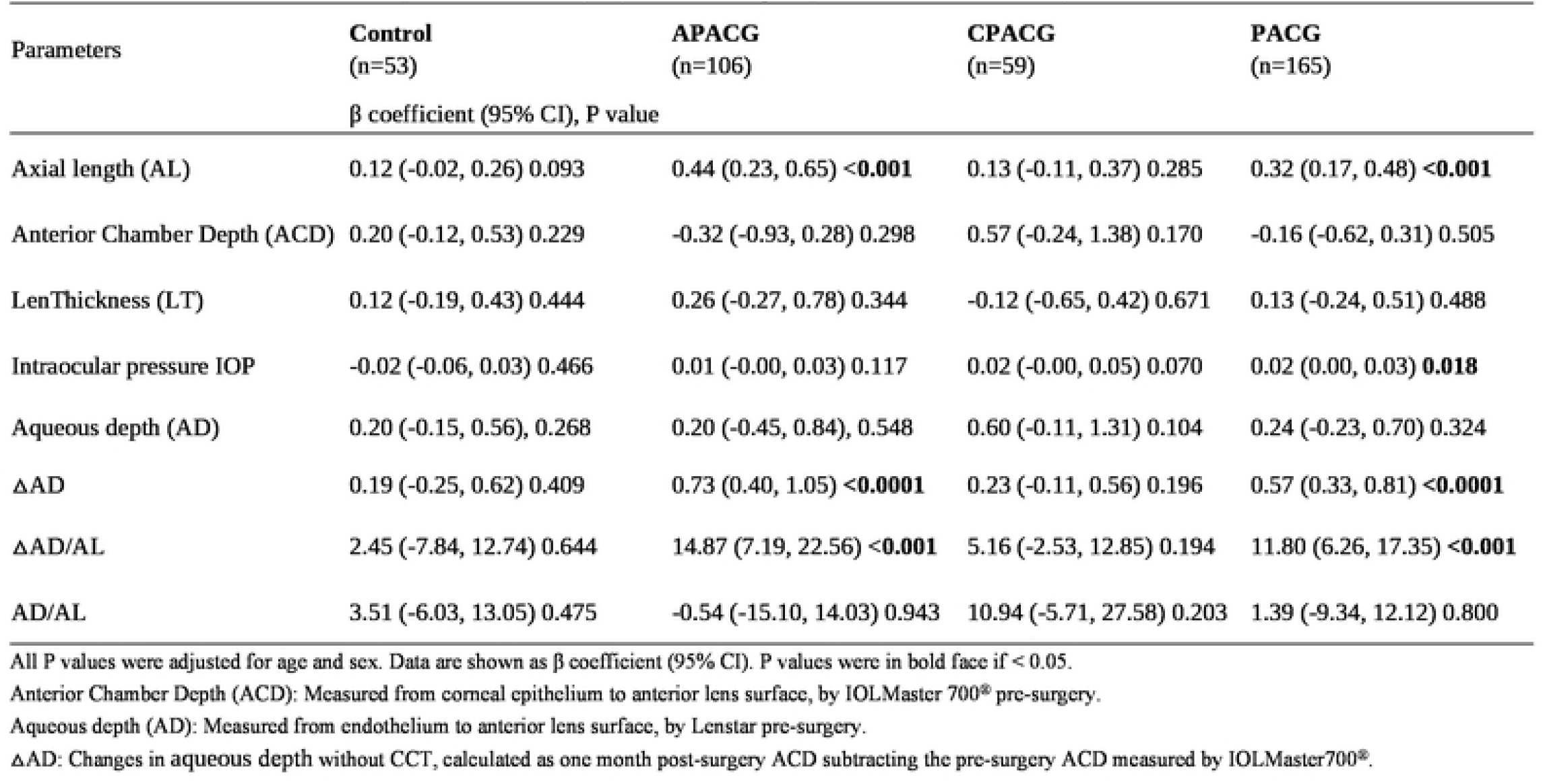
Risk factors associated with prediction error (PE) in different groups in multivariable analysis.

| Parameters | Control<br>(n=53) | APACG<br>(n=106) | CPACG<br>(n=59) | PACG<br>(n=165) |
| --- | --- | --- | --- | --- |
| | $\beta$ coefficient (95% CI), P value | | | |
| Axial length (AL) | 0.12 (-0.02, 0.26) 0.093 | 0.44 (0.23, 0.65) <0.001 | 0.13 (-0.11, 0.37) 0.285 | 0.32 (0.17, 0.48) <0.001 |
| Anterior Chamber Depth (ACD) | 0.20 (-0.12, 0.53) 0.229 | -0.32 (-0.93, 0.28) 0.298 | 0.57 (-0.24, 1.38) 0.170 | -0.16 (-0.62, 0.31) 0.505 |
| LenThickness (LT) | 0.12 (-0.19, 0.43) 0.444 | 0.26 (-0.27, 0.78) 0.344 | -0.12 (-0.65, 0.42) 0.671 | 0.13 (-0.24, 0.51) 0.488 |
| Intraocular pressure IOP | -0.02 (-0.06, 0.03) 0.466 | 0.01 (-0.00, 0.03) 0.117 | 0.02 (-0.00, 0.05) 0.070 | 0.02 (0.00, 0.03) **0.018** |
| Aqueous depth (AD) | 0.20 (-0.15, 0.56), 0.268 | 0.20 (-0.45, 0.84), 0.548 | 0.60 (-0.11, 1.31) 0.104 | 0.24 (-0.23, 0.70) 0.324 |
| ΔAD | 0.19 (-0.25, 0.62) 0.409 | 0.73 (0.40, 1.05) <0.0001 | 0.23 (-0.11, 0.56) 0.196 | 0.57 (0.33, 0.81) <0.0001 |
| ΔAD/AL | 2.45 (-7.84, 12.74) 0.644 | 14.87 (7.19, 22.56) <0.001 | 5.16 (-2.53, 12.85) 0.194 | 11.80 (6.26, 17.35) <0.001 |
| AD/AL | 3.51 (-6.03, 13.05) 0.475 | -0.54 (-15.10, 14.03) 0.943 | 10.94 (-5.71, 27.58) 0.203 | 1.39 (-9.34, 12.12) 0.800 |

All P values were adjusted for age and sex. Data are shown as $\beta$ coefficient (95% CI). P values were in bold face if $< 0.05$ .
Anterior Chamber Depth (ACD): Measured from corneal epithelium to anterior lens surface, by IOLMaster 700® pre-surgery.
Aqueous depth (AD): Measured from endothelium to anterior lens surface, by Lenstar pre-surgery.
ΔAD: Changes in aqueous depth without CCT, calculated as one month post-surgery ACD subtracting the pre-surgery ACD measured by IOLMaster700®.

With adjustment for age, sex, AL, and LT, multivariable linear regression analysis demonstrated that △AD was significantly associated with PE in total PACG (β = 0.57, 95% C.I.: 0.30 – 0.84, *P* < 0.001) and APACG groups (β = 0.67, 95% C.I.: 0.29 – 1.04, *P* < 0.001; Supplementary table 1). Moreover, △AD was also significantly associated with PE in the PACG patients with AL ≥ 22 mm (β = 0.55, 95% C.I.: 0.32 – 0.78, *P* < 0.001; Supplementary table 2); no association was found in the PACG patients with AL < 22 mm (β = 0.46, 95% C.I.: -0.31 – 1.22, *P* = 0.250).

## Discussion

Results from this study demonstrated that (1) PACG group, especially APACG, had significantly smaller proportions of eyes within PE ± 0.50 D than the control group; (2) AL,△AD, and the ratio of △AD/AL were significantly associated with PE in the PACG group, especially APACG patients; (3)△AD was significantly associated with PE in the PACG patients with AL ≥ 22 mm. Collectively, this study identified the ocular biometric parameters influencing the predicted refractive outcomes in the IOL implantation for PACG patients.

Previous studies have demonstrated that 71 – 82% eyes across the entire ACD range achieve within ± 0.50 D of PE [14, 15]; yet, only 25 – 77% of eyes with shallow ACD achieve within ± 0.50 D of PE [16, 17, 18-20]. Consistently, this study found that 71.69% control subjects achieve within ± 0.50 D of PE. Achieving satisfactory refractive outcomes remains a significant challenge in cataract surgery for shallow anterior chamber eyes (ACD < 3 mm) [21]. Advances in pre-operative biometry and IOL power calculation formulas can significantly improve the refractive outcomes in cataract surgery [22, 23]. However, refractive PE remains high in the eyes with shallow anterior chamber as compared to the eyes with normal ACD [16, 24]. The inaccuracy in effective lens position prediction mainly contributes to the errors in IOL power calculation [25, 26]. Pre-operative ACD indicates the axial position of the natural lens, and its deviation from average can significantly influence post-operative refractive outcomes [17]. The eyes with shallow ACD are frequently accompanied by relatively short AL and thicker lens [27], which can affect the prediction of effective lens position. Previous studies compared the impact of ACD, AL, and LT on the accuracy of IOL calculation formulas in eyes with shallow ACD [21, 28, 29].

However, the APACG group in our study had a lower percentage of eyes within ±0.50 D of PE, whereas the CPACG group showed no difference compared to the control group. Nevertheless, both the APACG and CPACG patients had shallower ACD and AD than the control subjects. These indicated that shallow ACD or AD might not be an associated factor for PE, which could be reflected by the lack of association of ACD and AD with PE in all study groups. Instead of ACD and AD, we demonstrated that the AL, △AD, and the △AD/AL ratio were associated with PE in the APACG patients, but not in CPACG patients, indicating that the AL and ΔAD mainly contribute to the PE of IOL power calculation in the APACG patients. Even adjusted for AL and LT, △AD is still significantly associated with PE. Notably, the association of △AD with PE was only found in the PACG patients with AL ≥ 22 mm, indicating that, other than AL, △AD could be a different mechanism contributing to the PE in IOL power calculation. How △AD exactly contributed to the PE in IOL power calculation requires further investigations. Our results showed that PACG patients with AL ≥ 22 mm had higher accuracy prediction than those with AL < 22 mm with low MAE and MedAE, more eyes within PE ± 0.50 D, and significant differences in absolute of PE. Our subgroup analysis revealed that the PACG patients with AL ≥ 22 mm and AD > 2 mm had more eyes within PE ± 0.50 D than those with AD ≤ 2 mm. Similarly, the PACG patients with AL < 22 mm and AD ≤ 2 mm had more eyes within PE ± 0.50 D than those with AD > 2 mm. These results remind us to pay more attention to eyes with an imblance ration of the AL and AD for the IOL power calculation in PACG patients, which is our target focus on intraocular lens calculations in the future.

There were several limitations in this study. First, this study only adopted the Barrett Universal II formula as the IOL calculation formula. As different impacts of ACD, AL, and LT on the accuracy of different IOL calculation formulas in eyes with shallow ACD has been reported [21], future studies can evaluate the association of different ocular biometric parameters with refractive outcomes using different IOL calculation formulae.

Second, the analyses of ocular biometric parameter measurement were not subdivided into different brands/sizes of IOL as it has been reported different changes in ACD can be observed with different sizes of IOL [30].

In summary, this study revealed the association of AL and △AD with PE in IOL power calculation in Chinese PACG patients. Adjustments in the IOL power calcuation are needed to improve the refractive outcomes of PACG, especially the APACG patients.

## Data Availability

Data can be made available

## Author Contribution

XHS and LW were responsible for formulating and composing the protocol. YPY and LW were responsible for extracting and scrutinizing data, interpreting findings. YPY, LW, TKN, were responsible for reseach annalyses, writing the text, critically reviewed the manuscript. XHY and JYC participated in providing valuable input to the manuscript. All authors read and approved the final manuscript.

## Data Availability

Data can be made available upon reasonable request to the corresponding author.

## Consent for publication

Not applicable.

## Ethics Approval

This study adhered to the Helsinki Declaration and was approved by the Medical Ethics Council of the Eye and ENT Hospital, Fudan University (2025268). Written informed consent was obtained from each participant.

## References

1. Tham YC, Li X, Wong TY, Quigley HA, Aung T, Cheng CY. Global prevalence of glaucoma and projections of glaucoma burden through 2040: a systematic review and meta-analysis. Ophthalmology.2014;121:2081–90 10.1016/j.ophtha.2014.05.013 PMID:24974815

2. Gao F, Wang J, Chen J, Wang X, Chen Y, Sun X. Etiologies and clinical characteristics of young patients with angle-closure glaucoma: a 15-year single-center retrospective study. Graefes Arch Clin Exp Ophthalmol. 2021; 259, 2379–2387 10.1007/s00417-021-05172-6 PMID:33876278

3. Tamm ER. The trabecular meshwork outflow pathways: structural and functional aspects. Exp Eye Res. 2009;88:648–55 10.1016/j.exer.2009.02.007 PMID:19239914

4. Sun X, Dai Y, Chen Y, Yu DY, Cringle SJ, Chen J, et al. Primary angle closure glaucoma: What we know and what we don’t know. Prog Retin Eye Res. 2017;57:26–45 10.1016/j.preteyeres.2016.12.003 PMID:28039061

5. You S, Liang Z, Yang K, Zhang Y, Oatts J, Han Y, et al. Novel Discoveries of Anterior Segment Parameters in Fellow Eyes of Acute Primary Angle Closure and Chronic Primary Angle Closure Glaucoma. Invest Ophthalmol Vis Sci. 2021;62:6. 10.1167/iovs.62.14.6 PMID:34730791

6. American Academy of Ophthalmology Glaucoma Panel. Preferred practice pattern® guidelines. Primary angle closure. San Francisco, CA, USA: American Academy of Ophthalmology 2015.

7. Azuara-Blanco A, Burr J, Ramsay C, Cooper D, Foster PJ, Friedman DS, et al. EAGLE study group. Effectiveness of early lens extraction for the treatment of primary angle-closure glaucoma (EAGLE): a randomised controlled trial. Lancet .2016;388:1389–97. 10.1016/S0140-6736(16)30956-4 PMID: 27707497

8. Huang J, Huang C. Zonulopathy and Its Relation to Primary Angle Closure Disease: A Review. J Glaucoma. 2024;33:931–39. 10.1097/IJG.0000000000002385 PMID: 38573908

9. Lee TE, Yoo C, Kim YY. The effects of peripheral anterior synechiae on refractive outcomes after cataract surgery in eyes with primary angle-closure disease. Medicine (Baltimore). 2021;100:e24673 10.1097/MD.0000000000024673 PMID:33832065

10. Seo S, Lee CE, Kim YK, Lee SY, Jeoung JW, Park KH. Factors affecting refractive outcome after cataract surgery in primary angle-closure glaucoma. Clin Exp Ophthalmol 2016;44:693–700. 10.1111/ceo.12762 PMID: 27082207

11. Hou M, Ding Y, Liu L, Li J, Liu X, Wu M. Accuracy of intraocular lens power calculation in primary angle-closure disease: comparison of 7 formulas. Graefes Arch Clin Exp Ophthalmol 2021;259:3739–3747. 10.1007/s00417-021-05295-w PMID: 34258655

12. Day AC, Cooper D, Burr J, Foster PJ, Friedman DS, Gazzard G, et al. Azuara-Blanco A. Clear lens extraction for the management of primary angle closure glaucoma: surgical technique and refractive outcomes in the EAGLE cohort. Br J Ophthalmol. 2018;102:1658–62. 10.1136/bjophthalmol-2017-311447 PMID:29453222

13. Huang H, Gao F, Sun X, Chen Y. Swept-source optical coherence tomography assessment of Schlemm’s canal after phacoemulsification with goniosynechialysis in Chinese patients with primary angle-closure glaucoma. Quant Imaging Med Surg. 2024;14:8119–30. 10.21037/qims-24-269 PMID:39698699

14. Melles RB, Holladay JT, Chang WJ. Accuracy of Intraocular Lens Calculation Formulas. Ophthalmology. 2018;125:169–78. 10.1016/j.ophtha.2018.02.008 PMID: 29784098

15. Melles RB, Kane JX, Olsen T, Chang WJ. Update on Intraocular Lens Calculation Formulas. Ophthalmology. 2019;126:1334–35. 10.1016/j.ophtha.2019.04.011 PMID: 30980854

16. Hipólito-Fernandes D, Luís ME, Serras-Pereira R, Gil P, Maduro V, Feijão J, Alves N. Anterior chamber depth, lens thickness and intraocular lens calculation formula accuracy: nine formulas comparison. Br J Ophthalmol. 2022;106:349–55. 10.1136/bjophthalmol-2020-317822 PMID: 33229347

17. Yan C, Yao K. Effect of Lens Vault on the Accuracy of Intraocular Lens Calculation Formulas in Shallow Anterior Chamber Eyes. Am J Ophthalmol. 2022;233:57–67. 10.1016/j.ajo.2021.07.011 PMID: 34293335

18. Eom Y, Kang SY, Song JS, Kim YY, Kim HM. Comparison of Hoffer Q and Haigis formulae for intraocular lens power calculation according to the anterior chamber depth in short eyes. Am J Ophthalmol. 2014;157:818–24. 10.1016/j.ajo.2013.12.017 PMID: 24345318

19. Yang S, Whang WJ, Joo CK. Effect of anterior chamber depth on the choice of intraocular lens calculation formula. PLoS One. 2017;12:e0189868. 10.1371/journal.pone.0189868 PMID: 29253884

20. Mo E, Lin L, Wang J, Huo Q, Yang Q, Liu E, et al. Clinical Accuracy of 6 Intraocular Lens Power Calculation Formulas in Elongated Eyes, According to Anterior Chamber Depth. Am J Ophthalmol. 2022;233:153–62. 10.1016/j.ajo.2021.07.017 PMID: 34303685

21. Yuan H, Zhang J, Han X, Ye J, Huang Y, Huang R, et al. Accuracy of 11 intraocular lens calculation formulas in shallow anterior chamber eyes. Acta Ophthalmol. 2024;102:e705–11.

22. Darcy K, Gunn D, Tavassoli S, Sparrow J, Kane JX, et al. Assessment of the accuracy of new and updated intraocular lens power calculation formulas in 10930 eyes from the UK National Health Service. J Cataract Refract Surg. 2020;46:2–7. 10.1016/j.jcrs.2019.08.014 PMID:32050225

23. Shrivastava AK, Nayak S, Mahobia A, Anto M, Pandey P. Accuracy of intraocular lens power calculation formulae in short eyes: A systematic review and meta-analysis. Indian J Ophthalmol. 2022;70:740–8. 10.4103/ijo.IJO_934_21 PMID:35225507

24. Lee Y, Kim MK, Oh JY, Choi HJ, Yoon CH. Intraocular lens power calculation in eyes with a shallow anterior chamber depth and normal axial length. PLoS One. 2023;18:0288554. 10.1371/journal.pone.0288554 PMID: 37498877

25. Norrby S. Sources of error in intraocular lens power calculation. J Cataract Refract Surg. 2008;34:368–76. 10.1016/j.jcrs.2007.10.031 PMID: 18299059

26. Kim YC, Sung MS, Heo H, Park SW. Anterior segment configuration as a predictive factor for refractive outcome after cataract surgery in patients with glaucoma. BMC Ophthalmol. 2016;16:179. 10.1186/s12886-016-0359-1 PMID: 27756264

27. Jivrajka R, Shammas MC, Boenzi T, Swearingen M, Shammas HJ. Variability of axial length, anterior chamber depth, and lens thickness in the cataractous eye. J Cataract Refract Surg. 2008;34:289–94. 10.1016/j.jcrs.2007.10.015 PMID: 18242456

28. Gao R, Zhang J, Han X, Huang Y, Huang R, Ye J, et al. Intraocular lens calculation formula selection for short eyes: based on axial length and anterior chamber depth. BMC Ophthalmol. 2025;25:21. 10.1186/s12886-024-03793-z PMID: 39819497

29. Wang Q, Jiang W, Lin T, Wu X, Lin H, Chen W. Meta-analysis of accuracy of intraocular lens power calculation formulas in short eyes. Clin Exp Ophthalmol. 2018;46:356–63. 10.1111/ceo.13058 PMID: 28887901

30. Takahashi Y, Hirano T, Nakamura M, Chiku Y, Hoshiyama K, Akahane S, et al. Temporal Change in Anterior Chamber Depth after Combined Vitrectomy and Cataract Surgery Using Different Sizes of Intraocular Lens. J Clin Med. 2022;11:6430. 10.3390/jcm11216430 PMID: 36362658

